# *Blastocystis* in the cervix of Polish women: a subtype found in a parenteral location is different to that found in the anus

**DOI:** 10.1101/2024.07.15.24310455

**Authors:** Barbara Suchońska, Adam Kaczmarek, Maria Wesołowska, Daniel Młocicki, Rusłan Sałamatin

## Abstract

**Objective:** The presence of *Blastocystis* spp. in a parenteral location—in the female genital tract—has been reported three times. The genetic material of the protozoan has been identified only once.

**Methods:** *Blastocystis* DNA was detected using real-time PCR.

**Results:** Thirty patients with so-called cervical erosions were examined. The presence of *Blastocystis* genetic material was confirmed in nine women. The authors are the first to confirm and identify the DNA of *Blastocystis* subtype ST1, ST6, and ST7 in samples taken from the ectocervix and the distal part of the cervical canal of women with large, symptomatic glandular ectopies which were resistant to standard treatment. In one case, in material from the cervix we identified a *Blastocystis* subtype which was different to that found in the anus of the same woman.

**Conclusions:** Our findings indicate that the presence of *Blastocystis* in the cervix is not, in any obvious way, associated with hygienic issues or neglect, but could be the result of women having vaginal intercourse with heterosexual men in whose semen this protozoan occurs. The possibility of *Blastocystis* occurrence in semen has been confirmed by recent publications as well as our own unpublished results. This discovery gives hope for the eradication of these organisms and thus to curing patients with chronic gynaecological problems.

## Introduction

*Blastocystis* spp. are anaerobic protozoans which commonly occur in the human digestive tract (El Safadi et al. 2014; Scanlan 2012; Turkeltaub et al. 2015; Zierdt 1991). They are characterized by considerable morphological as well as genetic polymorphism—28 distinct subtypes have been described so far (Tan 2008; Villalobos et al. 2014).

They are among the group of parasites which are most commonly detected in faecal samples, but there is considerable controversy as to their pathogenicity (Jimenez-Gonzalez et al. 2012; Scanlan 2012; Turkeltaub et al. 2015; Yakoob et al. 2010). There is no agreement on the role of *Blastocystis* in the digestive tract (Leder et al. 2005). However, it seems that outside the digestive tract, in environments which are untypical for this organism, the protozoan is a parasite (Escutia-Guzman et al. 2020; Wołynska and Soroczan 1972).

It is currently known that *Blastocystis* spp. do not occur in the vagina or the cervix under physiological conditions. It certainly is not part of the saprophytic flora of the female reproductive organ. Indeed, up until recent times, this location was not checked for their presence. Prior to 2022, only one report was published, in Polish, by Wołynska & Soroczan ‘[*Blastocystis hominis* Brumpt, 1912, (Phycomycetes) in the female genital tract]’, which described the presence of *Blastocystis* in vaginal swabs in patients with ‘erosions’ (Wołynska and Soroczan 1972).

Recently, two papers have appeared confirming the presence of *Blastocystis* in the cervix and vagina on the basis of a microscopic examination and genetic analysis (Escutia-Guzman et al. 2020; Villalobos et al. 2022). In research by Villalobos *et al*. the presence of this protozoan was described both in the vagina of women and in the semen of men infected with *Trichomonas vaginalis*. The Villalobos *et al*. (Villalobos et al. 2022) paper sheds new light on the mode of *Blastocystis* transmission. So far, it has been assumed that many parasitic infections can be transferred sexually, including those caused by *Entamoeba histolytica, T. vaginalis*, and *Toxoplasma gondii*: (Crespillo-Andujar et al. 2018). All of these may also be responsible for male infertility. Even though, in 2018 (Crespillo-Andujar et al. 2018), the possibility of *Blastocystis* infection being transmitted by sexual contact was not considered, we now know that this mode of transmission is possible (Villalobos et al. 2022).

In our paper, we present the occurrence of *Blastocystis* in an unusual location in the human body—in the cervix. The study attempted to find the route by which the infection took place, therefore, the presence of *Blastocystis* in the anuses of these women was also investigated.

## Material and Methods

### Study group and control group

The research was performed on patients reporting to the Cervical Counselling section of the Outpatient Clinic at the First Department of Obstetrics and Gynaecology; they were recruited for the research during routine visits to the clinic. Women were selected who were not menstruating and who had not had sexual intercourse within the last 24 hours, at the time the samples were taken.

Included in the study group were 30 regularly menstruating patients of reproductive age (18–50 years old), who had had so-called cervical erosion for at least one year, that is, an extensive glandular ectopy on the ectocervix, for which attempts at pharmacological treatment had been made. The patients qualified for this group were not pregnant, did not suffer from DM (diabetes mellitus), were not being treated with immunosuppressants, were not taking steroids (which lowers the immune response), and had not used local or systemic metronidazole or cotrimoxazole for the last three months. The control group consisted of 30 healthy women of reproductive age who were without any macroscopic lesions of the cervix, without reported chronic diseases, and without erosions. All patients gave their informed consent for participation in the study. A condition for inclusion in the study was also a current, normal Pap smear result. The patients’ data were anonymized for the study.

The samples were transferred to an employee of the Chair of Biology and Parasitology of Warsaw Medical University and the analysis itself was performed in the Department of Parasitology and Vector Borne Diseases of the National Institute of Hygiene where the material was stored, analysed, and utilized.

Cervical and anal swabs were taken from patients from the study group. Only cervical swabs were taken from patients from the control group.

### Sample collection

In the study group samples were collected by means of two separate dry swabs: one from the ectocervix and from the area of external os of the cervical canal; and a second from the anus at a minimum depth of 1 cm and collecting a minimal amount of the faecal material. For the control group, the sampling was limited to cervical swabs. Test tubes containing the material were transferred to the microbiology lab shortly after collection (Kaczmarek et al. 2022).

### Molecular identification and sequencing

DNA was isolated from the samples using Genomic Mini kits (A&A Biotechnology, Gdynia, Poland). A fragment of the small subunit ribosomal RNA (rRNA) was amplified using Bl18SPPF1 and BL18SR2PP primers (Poirier et al. 2011). Further analysis (purifying PCR products, sequencing, phylogenetic analysis) was performed as previously described by us (Kaczmarek et al. 2021). The sequences have been deposited with GenBank (accession numbers: XXXXX–XXXXX). The *Blastocystis* subtype nomenclature is according to Stensvold *et al*. (Stensvold et al. 2007).

This study was approved by the Bioethical Committee of Medical University of Warsaw: consent number KB1/175/20019.

## Results

### The group of patients with cervical erosion

Molecular analyses showed that *Blastocystis* DNA was present in cervical swabs from six patients and in anal swabs from four patients (Table 1).

**Table 1.**
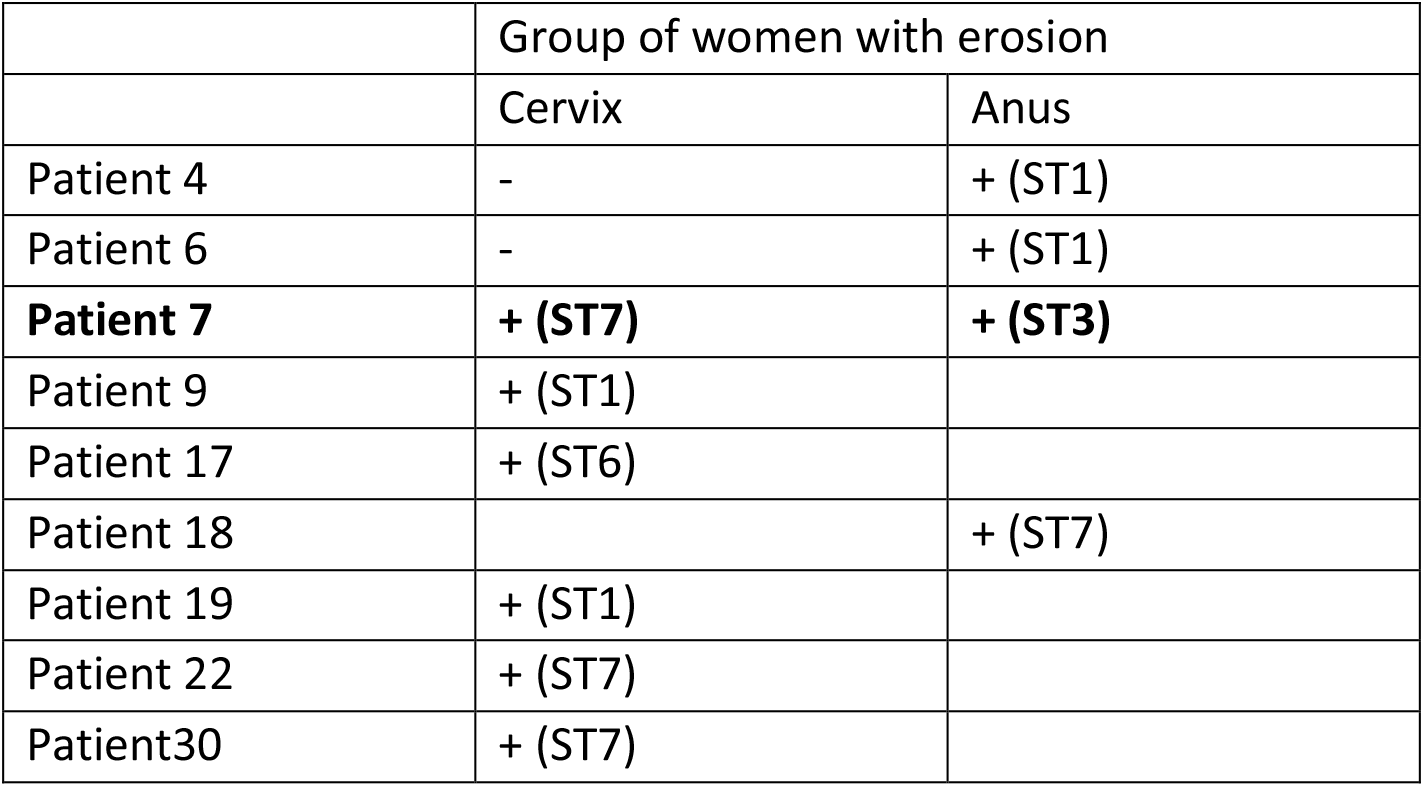
Blastocystis presence in swabs taken from the cervix and the anus

|  | Group of women with erosion |  |
| --- | --- | --- |
|  | Cervix | Anus |
| Patient 4 | - | + (ST1) |
| Patient 6 | - | + (ST1) |
| **Patient 7** | **+ (ST7)** | **+ (ST3)** |
| Patient 9 | + (ST1) |  |
| Patient 17 | + (ST6) |  |
| Patient 18 |  | + (ST7) |
| Patient 19 | + (ST1) |  |
| Patient 22 | + (ST7) |  |
| Patient30 | + (ST7) |  |

### Control group

*Blastocystis* was not detected in any control group patient.

## Discussion

Until recently, observations concerning the presence of *Blastocystis* in patients with cervical inflammation have been confirmed by only a few authors.

For a long time, the presence of the protozoan was detected on the basis of examining a microscopic preparation performed from an appropriate swab (Crespillo-Andujar et al. 2018; Engberts et al. 2007; Patino et al. 2008; Wołynska and Soroczan 1972). This method was also used by the authors of a paper from 2020 to confirm *Blastocystis* presence in the cervix of the described patient—this was photographically documented (Escutia-Guzman et al. 2020). Wołynska & Soroczan (Wołynska and Soroczan 1972) hypothesised that these microorganisms could be the etiological factor for non-healing erosions, which have been a therapeutic problem for gynaecologists. The suggestion was that the direct cause could be chronic hygienic neglect, which would justify the more common occurrence of these lesions in women over 30 years of age. The presence of this microorganism in 11.5% (47/312) of the analysed population was accompanied by colpitis in 51% (24/47) and cervical erosion in 32% (15/47) of the studied women. No pathology was found in the genital tract in 17% (8/47), which, according to the authors, was probably due to recent infection in this last subgroup. The analysed group was quite large, and encompassed 312 Polish women. Until recently this was the only paper concerning parenteral localization of *Blastocystis* and is still the only paper with such a large group of subjects, on the basis of which the authors attempted to draw clinical conclusions.

After 50 years, contemporary medicine has finally acquired many new diagnostic and therapeutic tools, which has been due to intensive developments in basic science. Currently, the presence of *Blastocystis* can be confirmed both by microscopic methods and those based on sequencing the genetic material of microorganisms. Thanks to the PCR analysis method, the diagnosis can be unequivocal and independent of the subjective opinion of the researcher (Edgar 2004; Feine et al. 2017; Kumar et al. 2018; Santín et al. 2011; Sepp et al. 1994). However, not all researchers use molecular tools, while some are unable to find an appropriate research group, which is confirmed by the latest reports (Crespillo-Andujar et al. 2018; Villalobos et al. 2022).

In 2020, a case of the extraintestinal occurrence of *Blastocystis* spp. was published concerning a patient with relatively mild symptoms of cervical inflammation (Escutia-Guzman et al. 2020). During a routine gynaecological visit, a cytological swab was taken in which the presence of *Blastocystis* sp. was noted, which was confirmed by PCR analysis of material taken from the anus (the PCR was not performed on the cervical material). It should not, however, be assumed that the *Blastocystis* present in the anus will be identical to that from the genital tract. It is worth considering that the cytological diagnostician evaluating the sample for oncological purposes was able to identify a *Blastocystis* infection in a very unlikely location, even though he was not an experienced parasitologist. The presence of *Blastocystis* infection is not routinely assessed in cytological smears and no appropriate standards have been implemented for this identification. That is why the above article deserves special attention. The authors stated that it was probably due to the topical use of prescription-free drugs that it was not possible to prove the presence of the protozoan in the cervix. This may be evidence of the effectiveness of metronidazole in the eradication of the infection, but could also mean that the cervix of the patient was not colonized. It seems that a clinical case of the parenteral occurrence of *Blastocystis* spp. has been described without any real confirmation of this hypothesis. On the basis of our research, we believe that *Blastocystis* can be present in the cervix and we have incontestable evidence of this fact. Some of our patients had the infection only in the cervix, but others had infections exclusively in the anus or in both locations. It seems that the presence of *Blastocystis* in the anus does not, with any certainty, indicate its presence in the vagina and the cervix. Even if the protozoan is present in both locations, it does not have to be the same subtype. We confirmed the presence of subtypes ST1, ST6, and ST7 in the cervix, whereas subtypes ST1, ST3 and ST7 were present in the anus. In the patient for whom the protozoan was present in both the vagina and the anus, it was determined with absolute certainty that these were two different subtypes: ST7 and ST3, respectively. The confirmation of the presence of *Blastocystis* spp. in the cervix with a simultaneous absence of typical or pronounced clinical symptoms, as we found in our patients, could suggest the extraordinary ability of these microorganisms to occupy new niches and adapt to new conditions. Unfortunately, the confirmation of their presence is not equivalent to finding the etiological agent for all cervical erosions. It is not known how to effectively eliminate this infection in patients with such an atypical parasite location.

In general, the incidence of *Blastocystis* in humans is estimated to be from 10% of the population in developed countries to 100% in developing countries, which is undoubtedly related to the level of hygiene (Ramírez et al. 2016; El Safadi et al. 2014; Scanlan 2012; Turkeltaub et al. 2015; Villegas-Gómez et al. 2016; Zierdt 1991). As previously mentioned, the clinical significance of *Blastocystis* infection is still unclear and gives rise to controversies. According to some researchers, *Blastocystis* is found more frequently in healthy carriers than in persons with diseases of the digestive tract (Leder et al. 2005).

In the vagina and the distal half of the cervix, bacterial, viral, and fungal infections are common. However, in this area, protozoan infections are much less common—most frequently the infectious agent is *T. vaginalis*: (Escutia-Guzman et al. 2020; Villalobos et al. 2022). Thus, the colonizing of the cervix by *Blastocystis* spp. in a patient with exceptionally light symptoms (limited to itching) is particularly atypical.

The authors of the most recent paper were able to perform molecular identification of *Blastocystis* subtypes ST1–ST3 in samples taken from the reproductive organs of six women (21.4% of patients tested) as well as three men (42.8%) infected by *T. vaginalis*, the protozoan responsible for most non-viral sexually transmitted infections (Villalobos et al. 2022). The unpublished results of our studies confirm that *Blastocystis* occurs in the semen of symptomatic men during routine testing for the presence of bacteria, viruses, and non-*Blastocystis* protozoa.

In the cited paper the samples were collected over four years, from 2015 to 2019. The exclusion criteria encompassed antibiotic therapy within at least 10 days preceding the collection of the swab, the use of vaginal drugs or having intercourse during the last three days, menstruation, and inadequate personal hygiene on the day the sample was collected. The presence of *Blastocystis* subtypes ST1–ST3 was noted in the material collected from both women and men. According to the authors of the quoted paper, this confirms the great ability of these microorganisms to colonize all available niches. It may turn out that the existence of a viral infection or an infection due to another protozoan increases the susceptibility of the patient to *Blastocystis* infection. This type of coincidence, however, requires further investigation. The limitation of this study is the lack of faecal sample analysis and the study of deep anal swabs.

Similar to the results in our study, Wołyńska and Soroczan pointed out a possibility for *Blastocystis* spp. being present in the vagina and the anus, but not necessarily simultaneously in the same patients (Wołyńska and Soroczan 1972). All the studied women in whom the infection was confirmed had symptoms of inflammation or of cervical erosion. In practically all the published papers so far, the presence of infections was probably associated with the subjects’ poor personal hygiene. During microscopic examination carried out by Wołyńska and Soroczan (Wołyńska and Soroczan 1972), and Escutia-Guzman (Escutia-Guzman et al. 2020), they found the presence of vacuolar forms of *Blastocystis*. Some doubts have been voiced about the way in which *Blastocystis* reaches the cervix and colonizes this area. This may be, but does not have to be, associated with an intense sex life or, as suggested by other authors, with poor hygiene. It is well known that some bacteria routinely present in the anus are also observed in small amounts in the cervical microbiome. This is true for *Escherichia coli* or *Enterococcus faecalis*. The mechanism of autonomous microorganism transfer, or that induced by everyday hygiene activities carried out by the women themselves through transfer due to the close proximity of the anus and vagina, has been used for years in the oral supplementation of gynaecological probiotics. After oral ingestion lactobacilli travel through the alimentary canal to the anus, colonizing it, and are then found in the vagina, which is colonized after about 14 days from initiation of the treatment (Gholiof et al. 2022; Reid et al. 2004; Reid et al. 2001) This solution ensures long term action and blocks the migration of pathogens from the anus, which is important, especially for preventing a recurrence of the infection. Finally, *Lactobacillus* bacteria present in the anus are a reservoir for colonizing the vagina. The same mechanism, though, this time not considered to be beneficial, could play a role in the transfer of *Blastocystis* and the colonization of female genital organs.

Taking into consideration that the presence of *Blastocystis* has been confirmed in semen, the possibility of sexual transfer cannot be excluded, especially through vaginal intercourse, with no connection to hygiene neglect or anal sex.

Summing up; *Blastocystis* can occur not only in the alimentary tract and anus but also in other locations in the anogenital area. In our studies, *Blastocystis* sp. was found in the cervixes of six out of thirty (20%) patients who had extensive non-healing erosions, and in none of the control group, who had no erosions. On the basis of the only paper in the available scientific literature with a large study group, we expected to find *Blastocystis* in cervixes which were modified by erosions.

Therefore, through genetic analysis, we confirmed the results of research conducted by Wołyńska 50 years ago. We did not identify any common factor occurring in patients with coexisting erosions and *Blastocystis* infection which would distinguish them from the remaining patients with erosions.

In a similar way to other researchers working on *Blastocystis* spp., we cannot conclude whether, in this cervical location, these protozoans are commensals or parasites. So far, research has not been performed on a sufficiently large group of patients to evaluate the scale of the phenomenon and the real frequency of *Blastocystis* spp. co-occurrence with cervical lesions. In our pilot study, involving a relatively small group of patients, we wanted to obtain an answer to the question as to whether the cervix’s colonization by *Blastocystis* can be confirmed and whether it is more common in women with erosions. We obtained a positive answer to both these questions. The question of whether this should be treated and if so, how, is still to be addressed.

## Author contributions

*B.S*., *A.K*., *M.W*., *D.M*., and *R.S*. were involved in the study concept and design. B.S. drafted the manuscript together with R.S., which all other authors critically revised. All authors approve the current version for submission. R.S. is the guarantor of the study.

## Data availability

The dataset from this study is held securely in coded form at Medical University of Warsaw. Data-sharing agreements prohibit making the dataset publicly available. However, the data can be made available upon reasonable request to the corresponding author (RS) after obtaining the necessary ethical and data-sharing approvals.

## Acknowledgements

We extend our gratitude to Laurence Taylor for meticulously reviewing the article, guaranteeing its precision and lucidity.

